# Association of home and neighbourhood conditions with anxiety and depression symptoms during the COVID-19 lockdown: Findings from the ALSPAC study

**DOI:** 10.1101/2024.07.23.24310858

**Authors:** Connor Pinkney, James B. Kirkbride, Andrew Boyd, Stanley Zammit, Joanne B. Newbury

**Affiliations:** Somerset NHS Foundation Trust; PsyLife Group, Division of Psychiatry, UCL, London, UK; Population Health Sciences, Bristol Medical School, University of Bristol, UK; UK Longitudinal Linkage Collaboration, University of Bristol, Bristol, UK; MRC Centre for Neuropsychiatric Genetics and Genomics, School of Medicine, Cardiff University, Cardiff, UK

**Keywords:** ALSPAC, lockdown, COVID-19, anxiety, depression, home conditions, neighbourhood conditions

## Abstract

**Background:** The first COVID-19 lockdown in the UK was initially hailed as a great leveller. However, given that people were restricted to their homes and immediate neighbourhoods, there were stark inequalities in how different people experienced lockdown. Nevertheless, evidence on the associations of home and neighbourhood conditions in mental health during lockdown is sparce.

**Methods:** Using data from the Avon Longitudinal Study of Parents and Children (ALSPAC), a UK population-based cohort, we examined associations of home and neighbourhood conditions with anxiety and depression symptoms at two points during the first UK lockdown in 2020 (23/03/20-15/06/20). Questionnaires were sent to the ALSPAC cohort at two timepoints (T1: April; T2: May/June), including validated measures of mental health, and questions about current home conditions and behaviours, including access to nature, garden access, house type, and household composition. Neighbourhood conditions were obtained via a novel linkage, and included neighbourhood deprivation, population density, social fragmentation, and greenspace. Main associations were examined using linear regression. Potential confounders were identified using a directed acyclic graph and included ethnicity, family psychiatric history, maternal social class, financial difficulties before lockdown, and previous anxiety and depression at age 18.

**Findings:** At T1, reduced access to nature (B=1.06, 95% CI=0.68-1.45, p<0.001) and neighbourhood deprivation (B=0.25, 95% CI=0.02-0.48) were associated with anxiety. Furthermore, reduced access to nature (B=0.99, 95% CI=0.57-1.40, p<0.001), no garden access (B=0.62, 95% CI=0.04-1.20, p=0.037), living alone (B=1.53, 95% CI=0.63-2.43, p=0.001), and neighbourhood deprivation (B=0.27, 95% CI=0.02-0.52, p=0.033) were associated with depression. Associations were similar, but often weaker, at T2. For example, there was strong evidence of associations only for access nature with anxiety (B=0.74, 95% CI=0.25-1.23, p=0.004); and for access to nature (B=1.06, 95% CI=0.50-1.61, p=0.001) and living alone (B=1.19, 95% CI=0.25-2.13, p=0.013) with depression.

**Interpretation:** Disadvantaged home and neighbourhood conditions, especially reduced access to nature and neighbourhood deprivation, were associated with more anxiety and depression symptoms during the first UK lockdown. In the case of future pandemics, mitigation efforts should be tailored to reduce the burden on mental health for those most vulnerable. However, the causality of these observational findings is uncertain.

## Introduction

There is evidence that the mental health of the UK population deteriorated during the first wave of the SARS-CoV-2 (COVID-19) pandemic, with increased levels of anxiety (Kwong et al., 2021) and mental distress (Pierce et al., 2020) particularly among young adults. Given that poor mental health will be the world’s biggest health problem by 2030 (World Health Assembly, 2012), it is important that research identifies factors that contributed to the deterioration of mental health during this period. This is necessary so that mental health interventions can be targeted to those still suffering. Furthermore, with climate change making pandemics increasingly likely (Carlson et al., 2022), evidence is needed to tailor future pandemic mitigation measures (such as mobility restrictions where people were largely confined to their homes; known as “lockdowns” in the UK), so that the collateral damage to mental health can be reduced.

A large body of evidence suggests that home and neighbourhood conditions play a role in the development of mental health problems.^5^ Home conditions and behaviours such as overcrowding or living alone, house type (e.g., detached house versus apartment), garden use, and access to nature, might each support or compromise mental health. For instance, previous research has associated living alone with depression (Stahl et al., 2017), and having access to a garden with better wellbeing (de Bell et al., 2020). Likewise, our neighbourhoods (i.e., our geographically defined local community) afford physical and social resources and stressors that may support or compromise our mental health; with a large body of research highlighting roles of population density (Newbury et al., 2016; Vassos et al., 2015), neighbourhood deprivation (Kirkbride et al., 2014; Newbury et al., 2022), social fragmentation (Allardyce et al., 2005; Evans et al., 2004), and greenspace (Engemann et al., 2019) with a range of mental health problems. Neighbourhoods can also buffer people from the negative mental health impacts of stressful life events (Kingsbury et al., 2020).

The COVID-19 pandemic, first recorded in Wuhan, China in December 2019 and declared a pandemic by the World Health Organisation in March 2020,^6^ saw the enforcement of strict lockdowns around the world with the intention of stopping mass infection.^7^ In the UK, the first national lockdown was announced on 23 March 2020, with most restrictions lifting by 14 August 2020. This was the strictest of all UK lockdowns, including the widespread closure of shops, schools, and services; and instructions to stay at home at all times, other than for a limited set of reasons (e.g., food shopping, medical appointments, one hour per day exercise, travel for essential “key workers” employment categories). We hypothesized that home and neighbourhood conditions would be particularly important to mental health during the first national lockdown, given that people were spending prolonged periods of time at home and were mostly restricted to their property, with limited access to their immediate neighbourhoods; creating a natural, quasi-experimental condition.

A handful of studies have examined the roles of home and neighbourhood conditions in mental health during national lockdowns. There is evidence that access to blue-green spaces was protective for mental health during lockdowns in Europe (Pouso et al., 2021). Furthermore, living alone and lacking access to outdoor space was associated with deteriorations in mental health among young people in Denmark (Groot et al., 2022). There is also evidence that associations between neighbourhood conditions and mental health were amplified during the UK lockdown (Teo et al., 2021). However, studies to date have often been cross-sectional, lacked adequate controls for potential confounders, used crude measures of neighbourhood conditions and mental health, and focussed on only one or two exposures or outcomes.

We therefore aimed to advance understanding on this topic using data from the Avon Longitudinal Study of Parents and Children (ALSPAC), a large UK birth cohort which rapidly responded with COVID-19 questionnaires sent to participants at two timepoints during the first UK lockdown, collecting detailed information about mental health, home conditions, and behaviours. A novel linkage was performed to link detailed data on neighbourhood conditions including population density, neighbourhood deprivation, social fragmentation, and greenspace to residential addressed during lockdown. Our hypotheses were that disadvantaged home and neighbourhood conditions would be associated with higher anxiety and depression symptoms during lockdown; worsening mental health during lockdown; and that associations would be stronger during lockdown compared to equivalent pre-pandemic assessments.

## Methods

### Study design and participants

ALSPAC is a UK birth cohort study (Boyd et al., 2013; Fraser et al., 2013; Northstone et al., 2019). Pregnant women residing in Bristol and surrounding areas (Southwest England) with an expected delivery date between 1^st^ April 1991 and 31^st^ December 1992 were approached to take part in the study, with 14,541 women initially recruited. When children were approximately 7 years of age, an attempt was made to bolster the initial sample to include additional children who met the original eligibility criteria, leading to a total sample size of 14,901 babies alive at 1 year of age. The catchment area includes the city of Bristol (population ∼714,000 in 2024), small towns and villages; with a mix of urban, suburban and rural environments (Boyd et al., 2019). Study data from age 22 onwards were collected and managed using REDCap (Research Electronic Data Capture) (Harris et al., 2009). The study website contains details of all the data and a fully searchable data dictionary and variable search tool: http://www.bristol.ac.uk/alspac/researchers/our-data/. Ethical approval for the study was obtained from the ALSPAC Ethics and Law Committee and the Local Research Ethics Committees. Informed consent for the use of data collected via questionnaires and clinics was obtained from participants following the recommendations of the ALSPAC Ethics and Law Committee at the time. The study conformed to the principles set by the Declaration of Helsinki.

### COVID-19 Questionnaires

The present study includes only the “index children” from the ALSPAC cohort. Participants were sent questionnaires at two time-points during the first UK lockdown, in April (T1) and May/June (T2) 2020. Questionnaires were only sent to those participants who had opted-in to receiving electronic surveys; as lockdown meant ALSPAC staff could not administer postal questionnaires. Participation in the questionnaire was voluntary. The questionnaire included assessments of mental health, home conditions, and behaviours.

### Anxiety and depression

Anxiety and depression were assessed at both T1 and T2. Anxiety was assessed using the Generalised Anxiety Disorder 7-item (GAD-7) questionnaire, which enquires about the occurrence of 7 anxiety symptoms over the previous 2 weeks, with a maximum score of 21 (Spitzer et al., 2006). Depression was assessed using the Short Moods and Feelings Questionnaire (SMFQ), which enquires about the occurrence of 13 depression symptoms over the previous 2 weeks, with a maximum score of 26 (Costello & Angold, 1988). In addition to examining scores at T1 and T2, we derived symptoms change scores by subtracting T1 scores from T2 scores (i.e., positive scores indicate increased anxiety/depression symptoms over time, and negative scores indicated decreased anxiety/depression symptoms over time.

Furthermore, at age 24 (∼4 years pre-pandemic), diagnostic thresholds for anxiety and depression were measured using the Clinical Interview Schedule Revised (CIS-R) (Lewis et al., 1992), a self-administered computerized interview that gave ICD-10 diagnoses of GAD and moderate-severe depression. The reporting period was the past month.

### Home conditions and behaviours

At T1, participants were asked various questions about their life during lockdown, including home conditions and behaviours. In the present study, home conditions included garden access, house type, and household composition. Behaviours included patterns of accessing nature.

*Nature access* behaviour was assessed by asking “Since the official lockdown, has the amount that you visit green space changed (e.g., park, beach, woodland; not your garden)?” and categorised such that 0=stayed the same/increased and 1=decreased. Note that we refer to this measure as “nature access” (rather than greenspace access) to distinguish it from the neighbourhood greenspace variable.

*Garden access* was assessed by asking participants “Do you have access to a garden?” and categorised such that 0=private/shared garden and 1=no garden.

*House type* was measured by asking participants “What type of accommodation do you live in?”. Due to small numbers in some categories, responses were categorised such that 0=detached house and 1=semi-detached house/terrace house/flat/apartment/maisonette/room in someone else’s house.

*Household composition* was measured by asking participants “Who do you live with?” and categorised such that 0=living with at least 1 other person and 1=living alone.

### Neighbourhood conditions

Neighbourhood conditions including population density, neighbourhood deprivation, social fragmentation, and greenspace were linked to participants home addresses during lockdown via non-identifying methods. These same neighbourhood variables were also linked to home addresses ∼4 years earlier when participants were aged 24, to investigate pre-lockdown associations. For all neighbourhood variables, scores were standardised such that the mean=0 and the standard deviation=1 (i.e., (X-M)/SD) to allow comparison between the variables.

*Population density* was derived from 2011 census data and defined as persons per hectare (Solmi et al., 2020).

*Neighbourhood deprivation* was based on the 2015 Index of Multiple Deprivation (IMD) (DETR, 2000), which ranks 32,844 small areas (Lower Layer Super Output Areas) according to various markers of deprivation in the domains of income; employment; education, skills and training; health and disability; crime; barriers to housing and services; and living environments.

*Social fragmentation* was based on z-scoring and summing the following 2011 census data: percentage of people who moved in the last year; percentage of unmarried people aged 16 and over; percentage of single person households; and percentage of privately rented households (Solmi et al., 2020).

*Greenspace* was assessed based on the Normalized Difference Vegetation Index (NDVI), derived from satellite imagery, described in detail previously (Fuertes et al., 2020). The present study uses estimates of greenness within a 300-meter buffer zone around residential addresses, which is commonly used by the World Health Organization as an accessibility threshold (Annerstedt Van Den Bosch et al., 2016).

### Confounders

Confounders were identified using a directed-acyclic-graph (DAG) (Figure 1). The confounders identified were ethnicity, family psychiatric history, maternal social class, previous anxiety or depression, and financial issues. By adhering to the rules of DAGs, sex did not require adjustment. These confounders are described further below.

**Figure 1.**
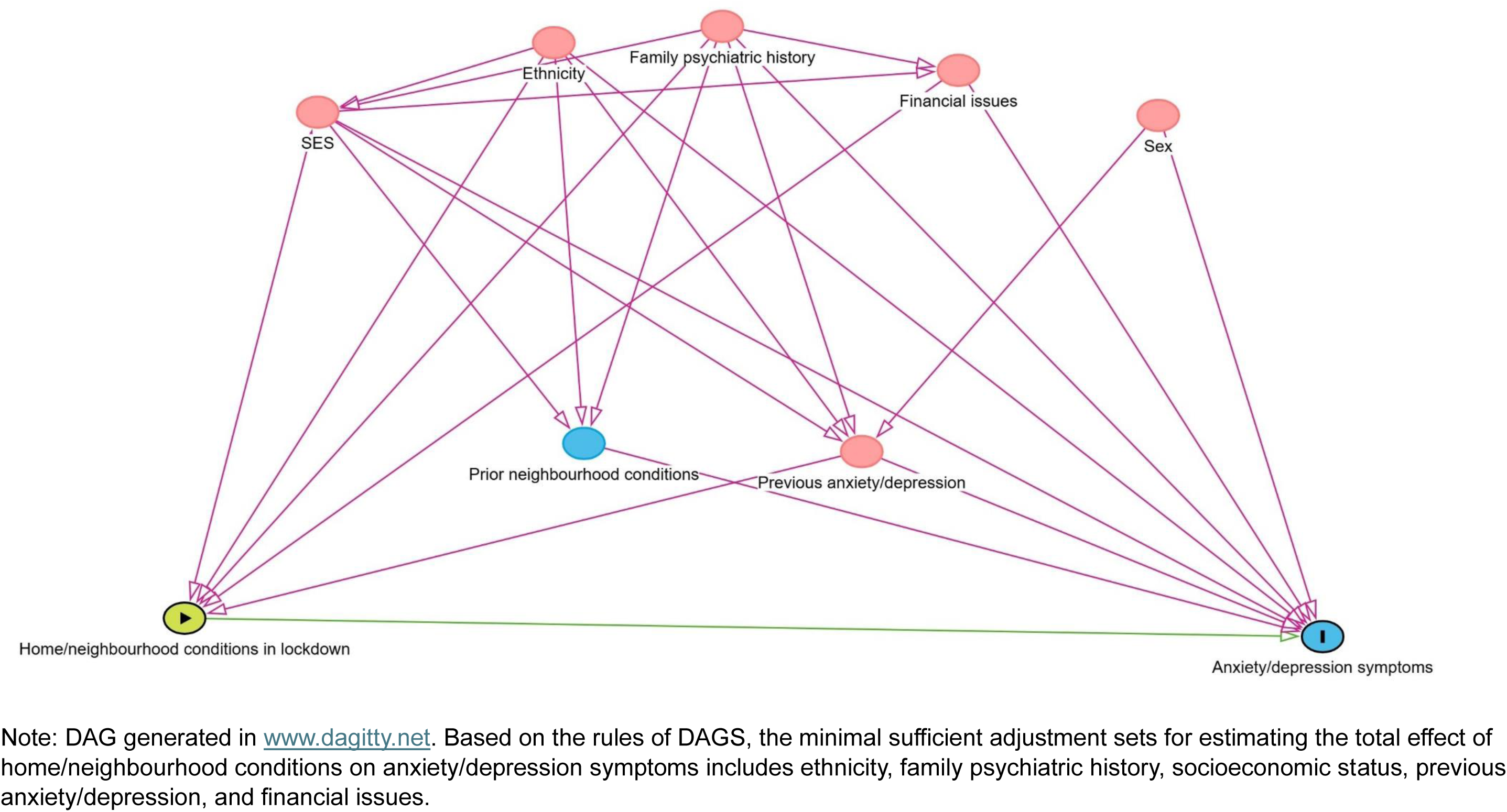
Directed acyclic graph identifying confounders of the associations of home and neighbourhood conditions with anxiety and depression

*Ethnicity* of the child was reported by mothers during pregnancy. Due to the small numbers within some ethnicities, ethnicity was dichotomized as White versus Non-White (all other ethnicities).

*Family psychiatric history* was reported by mothers and fathers during pregnancy, and defined as the presence of schizophrenia, depression, drug addiction, alcoholism, or any other psychiatric problem in the mother, father, or the mother/father’s biological mother or father.

*Maternal social class* was reported by mothers during pregnancy based on occupation. Occupations were defined as professional, managerial and technical, skilled non-manual, skilled manual, partly skilled, and unskilled.

*Previous anxiety and depression symptoms* were measured via face-to-face interviews at age 17 according to GAD-7 and the International Statistical Classification of Diseases and Related Health Problems ten (ICD criteria, respectively (Spitzer et al., 2006; World Health Organization, 1992)

*Financial issues* were assessed at the T2 COVID-19 questionnaire by asking participants how they managed financially before the pandemic (living comfortably/doing alright/just about getting by/finding it quite difficult/finding it very difficult).

### Statistical analysis

Statistical analyses were completed in Stata v18. First, characteristics of the sample who did versus did not respond to the COVID-19 questionnaires were described according to percentages, means, and standard deviations. Chi-square and t-tests were used to explore group differences. Second, linear regression models were used to examine the a) cross-sectional and b) longitudinal associations of home and neighbourhood conditions with anxiety and depression symptoms at a) T1 and b) T2, respectively. For each step, we first ran an unadjusted model, followed by a model adjusted for covariates.

All analyses were conducted following multiple imputation via chained equations and limited to those living in the UK who had responded to either COVID-19 questionnaires and had anxiety and depression data at any point. We imputed missing values for all mental health variables, home conditions variables, neighbourhood conditions variables, and covariates with missing data within this sample, with each variable specified as a continuous, ordinal, count, or binary variable, as appropriate. We imputed 5 datasets using a random seed. As a sensitivity analysis, we 1) repeated analyses for the change in anxiety and depression symptoms between T1 and T2; and 2) ran logistic regression models to examine the association of neighbourhood conditions at age 24 with diagnosed thresholds for ICD-10 anxiety and depression at age 24.

## Results

### Sample characteristics

The first COVID-19 questionnaire was sent to 5,848 ALSPAC participants, of whom 2,973 (50.8%) UK-based participants responded. The second COVID-19 questionnaire was sent to 6,141 ALSPAC participants, of whom 2,710 (44.1%) UK-based participants responded. The mean age of the participants was 28.3 years (range=27.3–29.3). We excluded respondents who were living abroad due to the study investigating lockdown in the UK. In total, 3,388 participants living in the UK responded to either questionnaire, forming the analytic sample.

Over two-thirds of respondents were female (female: N=2,392, 70.7%) and just under one-third were male (N=993; 29.3%). Most participants were ethnically White (N=2,923; 96.8%); with the remaining 97 respondents (3.2%) including Bangladeshi, Black African, Black Caribbean, Chinese, Indian, Pakistani, and other ethnicities.

Most participants were from higher socioeconomic groups, with N=2,206 (81.1%) of participants having had mothers with professional, managerial, technical, or skilled non-manual occupations. Respondents versus non-respondents were more likely to be female (*χ*^2^=261.8, p<0.001), White (*χ*^2^=11.8, p=0.001), and to have a family psychiatric history (*χ*^2^=7.1, p=0.008).

Nearly half of participants (N=1,189, 46.2%) reported living comfortably before the lockdown, with 72 (2.8%) finding it quite/very difficult. The average anxiety and depression symptom scores were M=6.2 (SD=5.3) and M=6.2 (SD=5.6) at T1, and M=6.1 (SD=5.3) and 6.6 (SD=5.7), respectively.

Nearly half of participants (N=1,323, 46.6%) reported having decreased access to nature since lockdown; 17.9% (N=512) did not have a garden; 58.8% (N=1,680) lived in a property other than detached house (e.g., semi-detached, terrace, flat, room in someone else’s house); and 5.8% (N=165) lived alone.

Mean population density was 39 people per hectare (SD=31); 28.0% (N=747) lived in deprived neighbourhoods; mean neighbourhood social fragmentation score was 0.6 (SD=3.9); and mean greenspace was NDVI=0.6 (SD=0.1) (NDVI values between 0.3 and 0.6 indicate areas with sparse vegetation cover; NDVI values between 0.6 and 0.9 indicate areas with dense and healthy vegetation cover).

### Association of home and neighbourhood conditions with anxiety and depression symptoms at T1

Association of home and neighbourhood conditions with anxiety and depression symptoms at T1 (∼April 2020) are shown in Table 2, with fully adjusted results shown in Figure 2.

**Figure 2.**
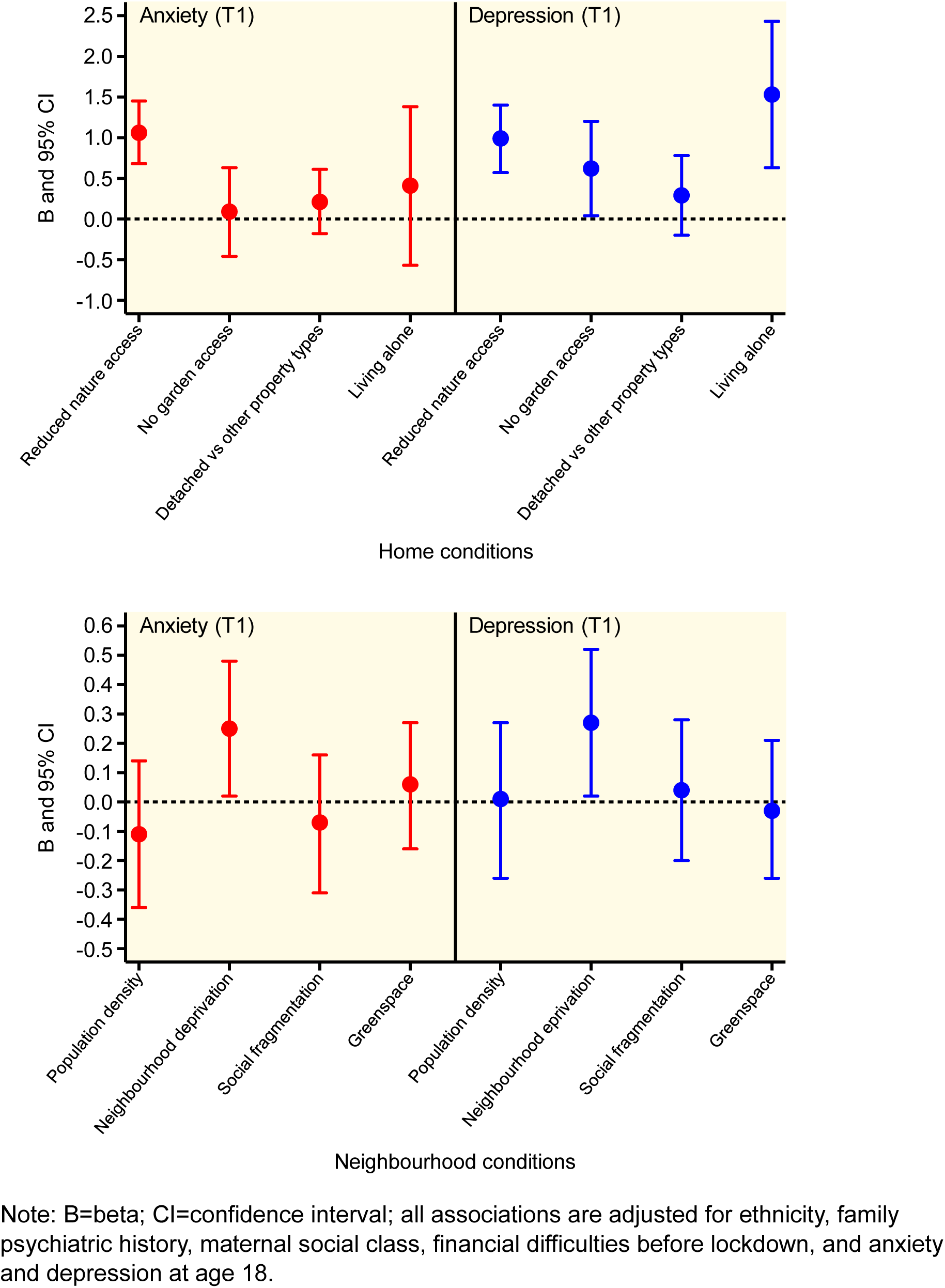
Adjusted associations of home and neighbourhood conditions with anxiety and depression at T1

**Table 1.** Sample characteristics for those who did versus did not respond to the COVID-19 questionnaires.

| Sample characteristics | Responded to one or both | | Responded to neither | | $\chi^2 / T$ | P-value |
| --- | --- | --- | --- | --- | --- | --- |
|  | N/M | %/SD | N/M | %/SD |  |  |
| Sex |  |  |  |  |  |  |
| Male | 993 | 29.3 | 1,232 | 50.2 |  |  |
| Female | 2,392 | 70.7 | 1,224 | 49.8 | 261.8 | <0.001 |
| Ethnicity |  |  |  |  |  |  |
| All other ethnicities <sup>a</sup> | 97 | 3.2 | 108 | 5.1 |  |  |
| White | 2,923 | 96.8 | 2,001 | 94.9 | 11.8 | 0.001 |
| Family psychiatric history |  |  |  |  |  |  |
| No | 1,081 | 35.8 | 815 | 39.5 |  |  |
| Yes | 1,938 | 64.2 | 1,250 | 60.5 | 7.1 | 0.008 |
| Maternal social class <sup>b</sup> |  |  |  |  |  |  |
| 1 – Professional | 143 | 5.3 | 95 | 5.0 |  |  |
| 2 – Managerial and technical | 951 | 34.9 | 627 | 33.0 |  |  |
| 3 – Skilled non-manual | 1,112 | 40.9 | 784 | 41.2 |  |  |
| 4 – Skilled manual | 101 | 3.7 | 67 | 3.5 |  |  |
| 5 – Partly skilled | 352 | 12.9 | 283 | 14.9 |  |  |
| 6 – Unskilled | 63 | 2.3 | 47 | 2.5 | 4.8 | 0.444 |
| Financial difficulties pre-lockdown |  |  |  |  |  |  |
| Living comfortably | 1,189 | 46.2 | - | - |  |  |
| Doing alright | 1,035 | 40.2 | - | - |  |  |
| Just about getting by | 278 | 10.8 | - | - |  |  |
| Finding it quite difficult | 60 | 2.3 | - | - |  |  |
| Finding it very difficult | 12 | 0.5 | - | - | - | - |
| Anxiety at age 18 |  |  |  |  |  |  |
| No | 2,026 | 91.3 | 1,198 | 93.01 |  |  |
| Yes | 193 | 8.7 | 90 | 6.99 | 3.2 | 0.073 |
| Depression at age 18 |  |  |  |  |  |  |
| No | 1,825 | 82.2 | 1,094 | 84.9 |  |  |
| Yes | 394 | 17.8 | 194 | 15.1 | 4.2 | 0.04 |
| Anxiety symptoms score |  |  |  |  |  |  |
| T1 | 6.2 | 5.3 | - | - |  |  |
| T2 | 6.1 | 5.3 | - | - | - | - |
| Depression symptoms score |  |  |  |  |  |  |
| T1 | 6.2 | 5.6 | - | - |  |  |
| T2 | 6.6 | 5.7 | - | - | - | - |
| Nature access |  |  |  |  |  |  |
| Same or increased | 1,517 | 53.4 | - | - | - | - |
| Decreased since lockdown | 1,323 | 46.6 | - | - | - | - |
| Garden access |  |  |  |  |  |  |
| Yes | 2,346 | 82.1 | - | - | - | - |
| No | 512 | 17.9 | - | - | - | - |
| Property type |  |  |  |  |  |  |
| Detached house | 1,178 | 41.2 | - | - | - | - |
| All other property types | 1,680 | 58.8 | - | - | - | - |
| Household composition |  |  |  |  |  |  |
| Live with others | 2,689 | 94.2 | - | - | - | - |
| Live alone | 165 | 5.8 | - | - | - | - |
| Population density <sup>c</sup> | 39 | 31 | - | - | - | - |
| Neighbourhood deprivation |  |  |  |  |  |  |
| 1 – least deprived | 678 | 25.5 |  |  |  |  |
| 2 | 708 | 26.6 |  |  |  |  |
| 3 | 531 | 19.9 |  |  |  |  |
| 4 | 480 | 18.0 |  |  |  |  |
| 5 – most deprived | 267 | 10.0 |  |  |  |  |
| Social fragmentation <sup>d</sup> | 0.6 | 3.9 | - | - | - | - |
| Greenspace <sup>e</sup> | 0.6 | 0.1 | - | - | - | - |

Note: M=mean; SD=standard deviation; T=t-test statistic; $\chi^2$ =Chi-square; <sup>a</sup> due to small numbers within most ethnicities, all ethnicities other than White were grouped. These ethnicities included Bangladeshi, Black African, Black Caribbean, Chinese, Indian, Pakistani, and other ethnicities; <sup>b</sup> based on maternal occupation; <sup>c</sup> unit is persons per hectare; <sup>d</sup> sum of z-scored census information on population turnover, unmarried people, single person households, and privately rented households; <sup>e</sup> unit is the Normalized Difference Vegetation Index: range -1 to 1.

**Table 2.**
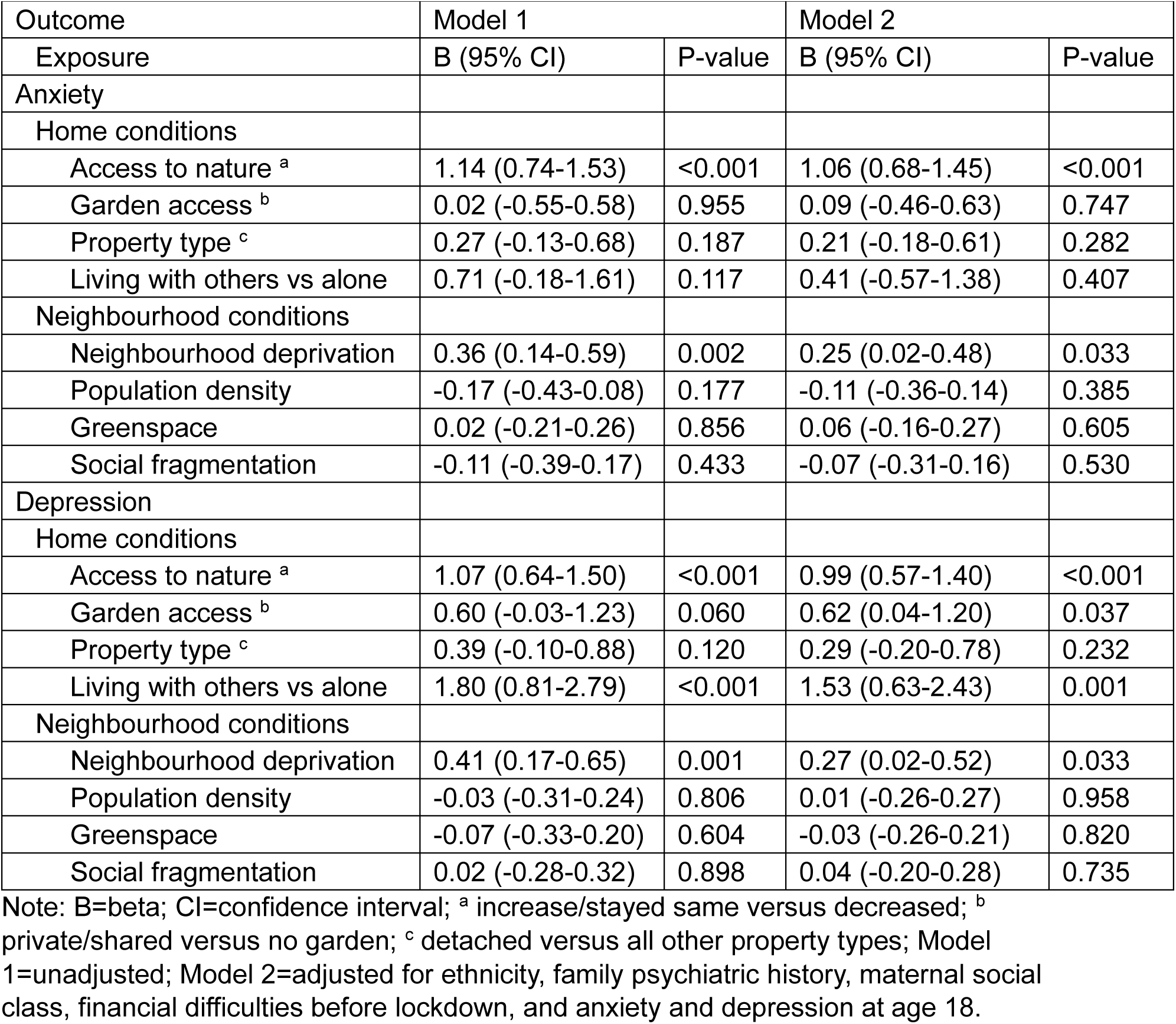
Associations of home and neighbourhood conditions with anxiety and depression symptoms at T1.

| Outcome | Model 1 |  | Model 2 |  |
| --- | --- | --- | --- | --- |
| Exposure | B (95% CI) | P-value | B (95% CI) | P-value |
| Anxiety |  |  |  |  |
| Home conditions |  |  |  |  |
| Access to nature <sup>a</sup> | 1.14 (0.74-1.53) | <0.001 | 1.06 (0.68-1.45) | <0.001 |
| Garden access <sup>b</sup> | 0.02 (-0.55-0.58) | 0.955 | 0.09 (-0.46-0.63) | 0.747 |
| Property type <sup>c</sup> | 0.27 (-0.13-0.68) | 0.187 | 0.21 (-0.18-0.61) | 0.282 |
| Living with others vs alone | 0.71 (-0.18-1.61) | 0.117 | 0.41 (-0.57-1.38) | 0.407 |
| Neighbourhood conditions |  |  |  |  |
| Neighbourhood deprivation | 0.36 (0.14-0.59) | 0.002 | 0.25 (0.02-0.48) | 0.033 |
| Population density | -0.17 (-0.43-0.08) | 0.177 | -0.11 (-0.36-0.14) | 0.385 |
| Greenspace | 0.02 (-0.21-0.26) | 0.856 | 0.06 (-0.16-0.27) | 0.605 |
| Social fragmentation | -0.11 (-0.39-0.17) | 0.433 | -0.07 (-0.31-0.16) | 0.530 |
| Depression |  |  |  |  |
| Home conditions |  |  |  |  |
| Access to nature <sup>a</sup> | 1.07 (0.64-1.50) | <0.001 | 0.99 (0.57-1.40) | <0.001 |
| Garden access <sup>b</sup> | 0.60 (-0.03-1.23) | 0.060 | 0.62 (0.04-1.20) | 0.037 |
| Property type <sup>c</sup> | 0.39 (-0.10-0.88) | 0.120 | 0.29 (-0.20-0.78) | 0.232 |
| Living with others vs alone | 1.80 (0.81-2.79) | <0.001 | 1.53 (0.63-2.43) | 0.001 |
| Neighbourhood conditions |  |  |  |  |
| Neighbourhood deprivation | 0.41 (0.17-0.65) | 0.001 | 0.27 (0.02-0.52) | 0.033 |
| Population density | -0.03 (-0.31-0.24) | 0.806 | 0.01 (-0.26-0.27) | 0.958 |
| Greenspace | -0.07 (-0.33-0.20) | 0.604 | -0.03 (-0.26-0.21) | 0.820 |
| Social fragmentation | 0.02 (-0.28-0.32) | 0.898 | 0.04 (-0.20-0.28) | 0.735 |

Note: B=beta; CI=confidence interval; <sup>a</sup> increase/stayed same versus decreased; <sup>b</sup> private/shared versus no garden; <sup>c</sup> detached versus all other property types; Model 1=unadjusted; Model 2=adjusted for ethnicity, family psychiatric history, maternal social class, financial difficulties before lockdown, and anxiety and depression at age 18.

After controlling for covariates, there was evidence that participants who reported reduced access to nature since the start of lockdown had more anxiety symptoms (B=1.06, 95% CI=0.68-1.45, p<0.001). Additionally, those living in more deprived neighbourhoods experienced more anxiety symptoms (B=0.25, 95% CI=0.02-0.48, p=0.033). No other home or neighbourhood conditions were associated with anxiety at T1.

Participants who reported reduced access to nature (B=0.99, 95% CI=0.57-1.40, p<0.001), had no garden (B=0.62, 95% CI=0.04-1.20, p=0.037), lived alone (B=1.53, 95% CI=0.63-2.43, p=0.001), and lived in more deprived neighbourhoods (B=0.27, 95 CI=0.02-0.52, p=0.033) also has more depression symptoms.

### Association of home and neighbourhood conditions with anxiety and depression symptoms at T2

Association of home and neighbourhood conditions with anxiety and depression symptoms at T1 (May/June 2020) are shown in Table 3, with fully adjusted results shown in Figure 3.

**Figure 3.**
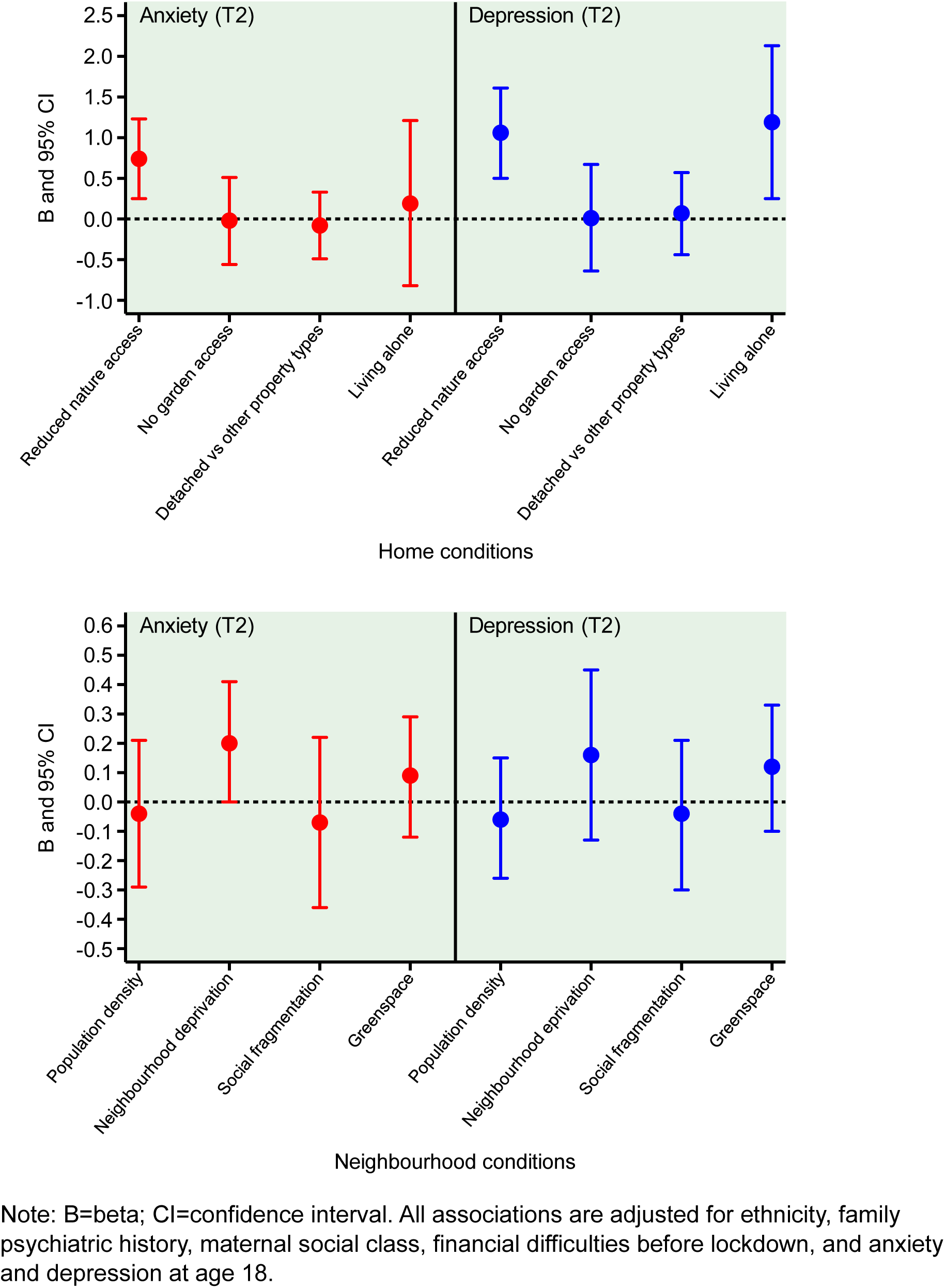
Adjusted associations of home and neighbourhood conditions with anxiety and depression at T2

**Table 3.** Associations of home and neighbourhood conditions with anxiety and depression symptoms at T2.

| Outcome | Model 1 |  | Model 2 |  |
| --- | --- | --- | --- | --- |
| Exposure | B (95% CI) | P-value | B (95% CI) | P-value |
| Anxiety |  |  |  |  |
| Home conditions |  |  |  |  |
| Access to nature | 0.82 (0.33-1.30) | 0.002 | 0.74 (0.25-1.23) | 0.004 |
| Garden access | -0.08 (-0.66-0.50) | 0.789 | -0.02 (-0.56-0.51) | 0.928 |
| Property type | -0.03 (-0.45-0.39) | 0.897 | -0.08 (-0.49-0.33) | 0.695 |
| Living with others vs alone | 0.50 (-0.43-1.43) | 0.287 | 0.19 (-0.82-1.21) | 0.704 |
| Neighbourhood conditions |  |  |  |  |
| Neighbourhood deprivation | 0.31 (0.11-0.51) | 0.003 | 0.20 (0.00-0.41) | 0.052 |
| Population density | -0.10 (-0.36-0.16) | 0.444 | -0.04 (-0.29-0.21) | 0.758 |
| Greenspace | 0.06 (-0.16-0.28) | 0.571 | 0.09 (-0.12-0.29) | 0.410 |
| Social fragmentation | -0.11 (-0.42-0.19) | 0.436 | -0.07 (-0.36-0.22) | 0.605 |
| Depression |  |  |  |  |
| Home conditions |  |  |  |  |
| Access to nature | 1.16 (0.59-1.73) | <0.001 | 1.06 (0.50-1.61) | 0.001 |
| Garden access | -0.00 (-0.70-0.69) | 0.992 | 0.01 (-0.64-0.67) | 0.969 |
| Property type | 0.14 (-0.38-0.66) | 0.589 | 0.07 (-0.44-0.57) | 0.793 |
| Household number | 1.53 (0.52-2.54) | 0.003 | 1.19 (0.25-2.13) | 0.013 |
| Neighbourhood conditions |  |  |  |  |
| Neighbourhood deprivation | 0.29 (0.01-0.57) | 0.040 | 0.16 (-0.13-0.45) | 0.264 |
| Population density | -0.11 (-0.33-0.12) | 0.361 | -0.06 (-0.26-0.15) | 0.602 |
| Greenspace | 0.09 (-0.14-0.32) | 0.457 | 0.12 (-0.10-0.33) | 0.288 |
| Social fragmentation | -0.09 (-0.37-0.19) | 0.523 | -0.04 (-0.30-0.21) | 0.743 |

Note: B=beta; CI=confidence interval; <sup>a</sup> increase/stayed same versus decreased; <sup>b</sup> private/shared versus no garden; <sup>c</sup> detached versus all other property types; Model 1=unadjusted; Model 2=adjusted for ethnicity, family psychiatric history, maternal social class, financial difficulties before lockdown, and anxiety and depression at age 18.

Again, reduced access to nature was associated with higher anxiety symptoms (B=0.74, 95% CI=0.25-1.23, p=0.004); though evidence became weaker for neighbourhood deprivation (B=0.20, 95% CI=0.00-0.41, p=0.052). Again, reduced access to nature (B=1.06, 95% CI=0.50-1.61, p=0.001) and living alone (B=1.19, 0.25-2.13, p=0.013) was associated with higher depression symptoms; but there was no longer evidence associating garden access or neighbourhood deprivation with depression symptoms.

### Association of home and neighbourhoods conditions with change in anxiety and depression symptoms between T1 and T2

Association of home and neighbourhood conditions with change in anxiety and depression symptoms between T1 and T2 are shown in Table 4. Interestingly, 12 out of 16 adjusted regression models were negative associations, suggesting that adverse home and neighbourhood conditions were associated with *decreasing* anxiety and depression symptoms between T1 and T2. However, confidence intervals were imprecise and included 0 across the board, providing no evidence against the null of there being no association.

**Table 4.** Association of home and neighbourhood conditions with change in anxiety and depression symptoms between T1 and T2.

| Outcome | Model 1 |  | Model 2 |  |
| --- | --- | --- | --- | --- |
| Exposure | B (95% CI) | P-value | B (95% CI) | P-value |
| Anxiety |  |  |  |  |
| Home conditions |  |  |  |  |
| Greenspace access <sup>a</sup> | -0.22 (-0.75-0.32) | 0.383 | -0.20 (-0.71-0.32) | 0.421 |
| Garden access <sup>b</sup> | -0.16 (-0.96-0.65) | 0.668 | -0.19 (-1.00-0.63) | 0.614 |
| Property type <sup>c</sup> | -0.31 (-0.82-0.20) | 0.212 | -0.31 (-0.83-0.21) | 0.218 |
| Living with others vs alone | -0.17 (-0.84-0.51) | 0.626 | -0.18 (-0.86-0.50) | 0.593 |
| Neighbourhood conditions |  |  |  |  |
| Neighbourhood deprivation | -0.07 (-0.24-0.10) | 0.419 | -0.05 (-0.21-0.12) | 0.551 |
| Population density | 0.11 (-0.07-0.30) | 0.223 | 0.11 (-0.09-0.30) | 0.262 |
| Greenspace | 0.08 (-0.13-0.28) | 0.435 | 0.06 (-0.15-0.27) | 0.553 |
| Social fragmentation | -0.00 (-0.21-0.21) | 0.997 | -0.01 (-0.23-0.21) | 0.899 |
| Depression |  |  |  |  |
| Home conditions |  |  |  |  |
| Greenspace access <sup>a</sup> | 0.14 (-0.29-0.57) | 0.512 | 0.10 (-0.33-0.53) | 0.626 |
| Garden access <sup>b</sup> | -0.51 (-1.03-0.01) | 0.053 | -0.51 (-1.05-0.03) | 0.064 |
| Property type <sup>c</sup> | -0.27 (-0.68-0.14) | 0.191 | -0.25 (-0.65-0.16) | 0.230 |
| Living with others vs alone | -0.21 (-0.99-0.57) | 0.592 | -0.30 (-1.06-0.46) | 0.439 |
| Neighbourhood conditions |  |  |  |  |
| Neighbourhood deprivation | -0.03 (-0.23-0.18) | 0.779 | -0.02 (-0.22-0.18) | 0.855 |
| Population density | -0.02 (-0.25-0.21) | 0.849 | -0.01 (-0.25-0.24) | 0.958 |
| Greenspace | 0.13 (-0.11-0.36) | 0.264 | 0.12 (-0.12-0.35) | 0.312 |
| Social fragmentation | -0.05 (-0.31-0.20) | 0.662 | -0.04 (-0.31-0.23) | 0.752 |

Note: B=beta; CI=confidence interval; <sup>a</sup> increase/stayed same versus decreased; <sup>b</sup> private/shared versus no garden; <sup>c</sup> detached versus all other property types. Model 1=unadjusted; Model 2=adjusted for ethnicity, family psychiatric history, maternal social class, financial difficulties before lockdown, and anxiety and depression at age 18.

### Association of neighbourhood conditions with anxiety and depression symptoms pre-lockdown (age 24)

Associations of neighbourhood conditions with anxiety and depression at age 24 (∼4 years pre-lockdown) are shown in Table 5 (note that equivalent home conditions were not available at age 24). Following covariate adjustment, there was no evidence associating neighbourhood deprivation or any other neighbourhood condition with anxiety or depression.

**Table 5.**
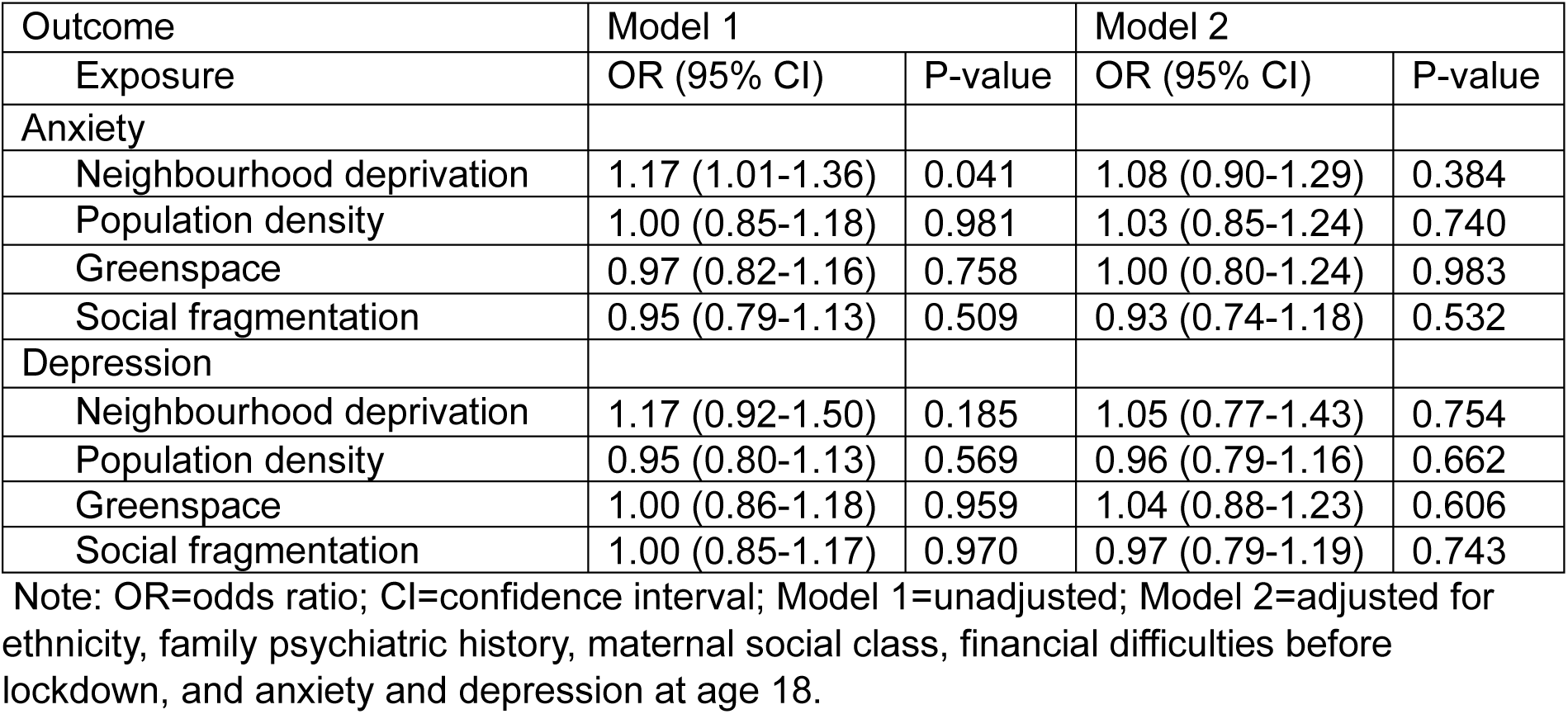
Association of neighbourhood conditions with anxiety and depression symptoms pre-lockdown (age 24)

## Discussion

This study explored the association of home and neighbourhood conditions with anxiety and depression symptoms in young adult participants of the ALSPAC study, at two timepoints during the first UK lockdown. Our hypotheses were that disadvantaged home and neighbourhood conditions would be associated with higher anxiety and depression symptoms in lockdown and worsening symptoms during lockdown; and that associations would be stronger compared to pre-pandemic equivalent measures.

We found evidence that home conditions (and behaviours) including reduced access to nature, having no garden, and living alone were associated with worse mental health symptoms; with reduced access to nature and living alone in particular associated with at least one additional depression symptom at both T1 and T2. We also found evidence associating neighbourhood deprivation with more anxiety and depression symptoms at T1, but not T2. No other neighbourhood conditions were associated with anxiety or depression. Additionally, there was an intriguing trend for disadvantaged home and neighbourhood conditions to be associated with decreasing symptoms over time, though there was no evidence against the null. Finally, we found little evidence associating neighbourhood conditions with anxiety and depression pre-pandemic at age 24. Thus, our hypotheses were only partly supported.

Our findings are consistent with a substantial body of evidence that exposure to nature/greenspace is beneficial for mental health (Houlden et al., 2018). Thus, restricted mobility during the first UK lockdown could have undermined mental health by reducing the therapeutic benefits of exposure to nature. Additionally, social isolation is a potent risk factor for psychopathology (Matthews et al., 2015), and its impact could have been amplified during lockdown. Finally, decades of research demonstrate patterning of mental health problems such as depression according to neighbourhood deprivation (Kirkbride et al., 2014).

However, against our hypotheses, there was no evidence that associations of home and neighbourhood conditions with anxiety and depression increased between T1 and T2. In fact, point estimates were often smaller at T2 versus T1; and disadvantaged home and neighbourhood conditions were often associated with decreasing symptoms over time (though evidence for this was weak). We note that the lockdown rules were strictest at the start of lockdown in April 2020. The relaxation of certain measures (e.g., being allowed to travel for exercise) in May/June 2020 would likely have enabled participants to benefit more from positive environments outside their home and immediate neighbourhoods, which may have given rise to reduced associations at T2 versus T1. Additionally, an unforeseen consequence of the pandemic was a change in living arrangements for many, with many young adults moving back to their family homes. For instance, 15% of 30 year-olds in the UK reported changes to their living arrangements by May 2020 because of COVID-19 (Evandrou et al., 2021). These societal changes may also have given rise to reduced associations at T2 versus T1.

### Strengths and limitations

This study has several strengths. It is one of only a handful of studies to investigate the association of home and neighbourhood conditions with mental health during lockdown. Due to its existing infrastructure, the ALSPAC team were able to rapidly respond to lockdown with detailed questionnaires capturing mental health at two timepoints. ALSPAC’s longitudinal design also allowed for the adjustment of key potential confounders collected prospectively, including measures of SES and prior mental health, which may have simultaneously led to disadvantaged home/neighbourhood conditions and mental health problems. Neighbourhood conditions were also obtained from a state-of-the-art linkage with objective data; yet being neighbourhood measures may suffer from ecological fallacy.

The study also has limitations. First, though the design was longitudinal, there were important differences in assessments pre-pandemic and during the pandemic which impeded certain analyses, such as direct comparisons of associations immediately before and at the start of lockdown. For instance, though we explored associations with prior mental health at age 24, this was limited to neighbourhood conditions, and based on different measures of anxiety and depression. Second, shared method variance may have influenced results, given that home conditions and mental health were both reported during the same questionnaires. For instance, being reminded of living alone during the questionnaire may have negatively impacted mood. Moreover, the strongest evidence was for self-reported nature access behaviours since lockdown, but these may have been subject to reverse causation. That is, people with existing mental health problems may have been more likely to decrease their access to nature. Indeed, it was notable that self-reported nature access, but not objectively measured greenspace, was associated with mental health. Third, the measures including garden access and property type would have missed key contextual information. For instance, neither the garden access nor property type measures captured the size or quality of the garden/house. The increasing sophistication of linkage to environmental and demographic data (e.g., land registry) and incorporation of satellite imagery opens new opportunities for more objective and detailed investigations to the roles of home and neighbourhood conditions in mental health. Fourth, responders were not representative of the general population (75% were female), and, overall, ALSPAC participants – reflecting the catchment area’s population characteristics at the time - are more affluent and less diverse than the UK population (Golding et al., 2001); thereby reducing the generalisability of the findings to other populations in the UK. Fifth, relatedly, participants were young adults. The extent that findings generalise to other age groups, including children and older adults, is uncertain. Finally, causality from observational findings remain uncertain.

## Conclusions

Our findings, based on the natural quasi-experiment created by the first UK lockdown, provide evidence that young adults living alone, with no garden, with reduced access to nature, and in deprived neighbourhoods had more anxiety and depression during lockdown. At the least, this highlights that mental health problems were experienced unequally in this population, with those most affected also disproportionately disadvantaged by lockdown. As climate change is increasing the likelihood of future pandemics, policymakers should urgently consider the cost-benefits of different crisis management approaches and whether mitigation efforts (e.g., lockdowns) can be tailored to individual circumstances to reduce impacts on mental health. However, our study has some limitations which impede a causal interpretation, including shared method variance, crude measures for some variables (e.g., garden access), inconsistent and temporally imprecise mental health measures, and uncertainties around address. Since the pandemic, there has been a shift in data science, with an increasingly sophisticated linkage capability that allows linkage of spatiotemporally precise and objective data on neighbourhood conditions, home conditions, meteorology, and mental health to large cohort studies with diverse sample characteristics and geographies (e.g., the UK Longitudinal Linkage Collaboration, which has partnered with 21 longitudinal cohorts in the UK). Moreover, the growth of wearable technologies and app-based mental health assessments greatly enhances future opportunity to explore the roles of home and neighbourhood conditions in mental health in real-time.

## Data Availability

All data produced in the present study are available upon reasonable request to the authors

## Corresponding author

Joanne B. Newbury, PhD, Population Health Sciences, Bristol Medical School, University of Bristol, Canynge Hall, Bristol, BS8 2PN, United Kingdom.

## Acknowledgements

We are extremely grateful to all the families who took part in this study, the midwives for their help in recruiting them, and the whole ALSPAC team, which includes interviewers, computer and laboratory technicians, clerical workers, research scientists, volunteers, managers, receptionists, and nurses.

## References

Allardyce J., Gilmour H., Atkinson J., Rapson T., Bishop J. and McCreadie R. (2005) Social fragmentation, deprivation and urbanicity: Relation to first-admission rates for psychoses. The British Journal of Psychiatry, 187, 401–406. doi:10.1192/bjp.187.5.401

Annerstedt Van Den Bosch M., Mudu P., Uscila V., Barrdahl M., Kulinkina A., Staatsen B.,… Egorov A. I. (2016) Development of an urban green space indicator and the public health rationale. Scandinavian Journal of Public Health, 44, 159–167. doi:10.1177/1403494815615444

Boyd A., Golding J., Macleod J., Lawlor D. A., Fraser A., Henderson J.,… Davey Smith G. (2013) Cohort profile: The ‘Children of the 90s’—the index offspring of the Avon Longitudinal Study of Parents and Children. International Journal of Epidemiology, 42, 111–127. doi:10.1093/ije/dys064

Boyd A., Thomas R., Hansell A. L., Gulliver J., Hicks L. M., Griggs R.,… Golding J. (2019) Data Resource Profile: The ALSPAC birth cohort as a platform to study the relationship of environment and health and social factors. International Journal of Epidemiology, 48, 1038–1039k. doi:10.1093/ije/dyz063

Carlson C. J., Albery G. F., Merow C., Trisos C. H., Zipfel C. M., Eskew E. A.,… Bansal S. (2022) Climate change increases cross-species viral transmission risk. Nature, 607, 555–562. doi:10.1038/s41586-022-04788-w

Costello E. J. and Angold A. (1988) Scales to assess child and adolescent depression: checklists, screens, and nets. Journal of the American Academy of Child and Adolescent Psychiatry, 27, 726–737. doi:10.1097/00004583-198811000-00011

de Bell S., White M., Griffiths A., Darlow A., Taylor T., Wheeler B. and Lovell R. (2020) Spending time in the garden is positively associated with health and wellbeing: Results from a national survey in England. Landscape and Urban Planning, 200, 103836. doi:10.1016/j.landurbplan.2020.103836

DETR. (2000). Indices of Deprivation 2000. Retrieved from London:

Engemann K., Pedersen C. B., Arge L., Tsirogiannis C., Mortensen P. B. and Svenning J.-C. (2019) Residential green space in childhood is associated with lower risk of psychiatric disorders from adolescence into adulthood. Proceedings of the National Academy of Sciences, 116, 5188–5193. doi:10.1073/pnas.1807504116

Evandrou M., Falkingham J., Qin M. and Vlachantoni A. (2021) Changing living arrangements and stress during COVID-19 lockdown: Evidence from four birth cohorts in the UK. SSM-population health, 13, 100761. doi:10.1016/j.ssmph.2021.100761

Evans J., Middleton N. and Gunnell D. (2004) Social fragmentation, severe mental illness and suicide. Social Psychiatry and Psychiatric Epidemiology, 39, 165–170. doi:10.1007/s00127-004-0733-9

Fraser A., Macdonald-Wallis C., Tilling K., Boyd A., Golding J., Davey Smith G.,… Ness A. (2013) Cohort profile: the Avon Longitudinal Study of Parents and Children: ALSPAC mothers cohort. International Journal of Epidemiology, 42, 97–110. doi:10.1093/ije/dys066

Fuertes E., Markevych I., Thomas R., Boyd A., Granell R., Mahmoud O.,… Henderson J. (2020) Residential greenspace and lung function up to 24 years of age: The ALSPAC birth cohort. Environment International, 140, 105749. doi:10.1016/j.envint.2020.105749

Golding, Pembrey and Team A. S. (2001) ALSPAC–The Avon Longitudinal Study of Parents and Children. Paediatric and Perinatal Epidemiology, 15, 74–87. doi:10.1046/j.1365-3016.2001.00325.x

Groot J., Keller A., Joensen A., Nguyen T.-L., Nybo Andersen A.-M. and Strandberg-Larsen K. (2022) Impact of housing conditions on changes in youth’s mental health following the initial national COVID-19 lockdown: A cohort study. Scientific Reports, 12, 1939. doi:10.1038/s41598-022-04909-5

Harris P. A., Taylor R., Thielke R., Payne J., Gonzalez N. and Conde J. G. (2009) Research electronic data capture (REDCap) - a metadata-driven methodology and workflow process for providing translational research informatics support. Journal of Biomedical Informatics, 42, 377–381. doi:10.1016/j.jbi.2008.08.010

Houlden V., Weich S., Porto de Albuquerque J., Jarvis S. and Rees K. (2018) The relationship between greenspace and the mental wellbeing of adults: A systematic review. PloS One, 13, e0203000. doi:10.1371/journal.pone.0203000

Kingsbury M., Clayborne Z., Colman I. and Kirkbride J. B. (2020) The protective effect of neighbourhood social cohesion on adolescent mental health following stressful life events. Psychological Medicine, 50, 1292–1299. doi:10.1017/S0033291719001235

Kirkbride J. B., Jones P. B., Ullrich S. and Coid J. W. (2014) Social deprivation, inequality, and the neighborhood-level incidence of psychotic syndromes in East London. Schizophrenia Bulletin, 40, 169–180. doi:10.1093/schbul/sbs151

Kwong A. S., Pearson R. M., Adams M. J., Northstone K., Tilling K., Smith D.,… Zammit S. (2021) Mental health before and during the COVID-19 pandemic in two longitudinal UK population cohorts. The British Journal of Psychiatry, 218, 334–343. doi:10.1192/bjp.2020.242

Lewis G., Pelosi A. J., Araya R. and Dunn G. (1992) Measuring psychiatric disorder in the community: a standardized assessment for use by lay interviewers. Psychological Medicine, 22, 465–486. doi:10.1017/S0033291700030415

Matthews T., Danese A., Wertz J., Ambler A., Kelly M., Diver A.,… Arseneault L. (2015) Social isolation and mental health at primary and secondary school entry: a longitudinal cohort study. Journal of the American Academy of Child and Adolescent Psychiatry, 54, 225–232. doi:10.1016/j.jaac.2014.12.008

Newbury J., Arseneault L., Caspi A., Moffitt T. E., Odgers C., Belsky D. W.,… Matthews T. (2022) Association between genetic and socioenvironmental risk for schizophrenia during upbringing in a UK longitudinal cohort. Psychological Medicine, 52, 1527–1537. doi:10.1017/S0033291720003347

Newbury J., Arseneault L., Caspi A., Moffitt T. E., Odgers C. L. and Fisher H. L. (2016) Why are children in urban neighborhoods at increased risk for psychotic symptoms? Findings from a UK longitudinal cohort study. Schizophrenia Bulletin, 42, 1372–1383. doi:10.1093/schbul/sbw052

Northstone K., Lewcock M., Groom A., Boyd A., Macleod J., Timpson N. and Wells N. (2019) The Avon Longitudinal Study of Parents and Children (ALSPAC): an update on the enrolled sample of index children in 2019. Wellcome Open Research, 4doi:10.12688/wellcomeopenres.15132.1

Pierce M., Hope H., Ford T., Hatch S., Hotopf M., John A.,… McManus S. (2020) Mental health before and during the COVID-19 pandemic: a longitudinal probability sample survey of the UK population. The Lancet Psychiatry, 7, 883–892. doi:10.1016/S2215-0366(20)30308-4

Pouso S., Borja Á., Fleming L. E., Gómez-Baggethun E., White M. P. and Uyarra M. C. (2021) Contact with blue-green spaces during the COVID-19 pandemic lockdown beneficial for mental health. Science of the Total Environment, 756, 143984. doi:10.1016/j.scitotenv.2020.143984

Solmi F., Lewis G., Zammit S. and Kirkbride J. B. (2020) Neighborhood characteristics at birth and positive and negative psychotic symptoms in adolescence: Findings from the ALSPAC birth cohort. Schizophrenia Bulletin, 46, 581–591. doi:10.1093/schbul/sbz049

Spitzer R. L., Kroenke K., Williams J. B. and Löwe B. (2006) A brief measure for assessing generalized anxiety disorder: the GAD-7. Archives of Internal Medicine, 166, 1092–1097. doi:10.1001/archinte.166.10.1092

Stahl S. T., Beach S. R., Musa D. and Schulz R. (2017) Living alone and depression: the modifying role of the perceived neighborhood environment. Aging & Mental Health, 21, 1065–1071. doi:10.1080/13607863.2016.1191060

Teo C., Kim C., Nielsen A., Young T., O’Campo P. and Chum A. (2021) Did the UK COVID-19 lockdown modify the influence of neighbourhood disorder on psychological distress? Evidence from a prospective cohort study. Frontiers in Psychiatry, 12, 702807. doi:10.3389/fpsyt.2021.702807

Vassos E., Agerbo E., Mors O. and Pedersen C. B. (2015) Urban–rural differences in incidence rates of psychiatric disorders in Denmark. The British Journal of Psychiatry, 208, 435–440. doi:10.1192/bjp.bp.114.161091

World Health Assembly. (2012). Global burden of mental disorders and the need for a comprehensive, coordinated response from health and social sectors at the country level: Report by the Secretariat. . Retrieved from Geneva: https://apps.who.int/iris/handle/10665/78898

World Health Organization. (1992). The ICD-10 classification of mental and behavioural disorders: Clinical descriptions and diagnostic guidelines. Geneva: World Health Organization.

